# Early transmission dynamics, spread, and genomic characterization of SARS-CoV-2 in Panama

**DOI:** 10.1101/2020.07.31.20160929

**Authors:** Danilo Franco, Claudia Gonzalez, Leyda E. Abrego, Jean Paul Carrera, Yamilka Diaz, Yaset Caisedo, Ambar Moreno, Oris Chavarria, Jessica Gondola, Marlene Castillo, Elimelec Valdespino, Melissa Gaitán, Jose Martínez-Mandiche, Lizbeth Hayer, Pablo Gonzalez, Carmen Lange, Yadira Molto, Dalis Mojica, Ruben Ramos, Maria Mastelari, Lizbeth Cerezo, Lourdes Moreno, Christl A. Donnelly, Nuno Rodrigues Faria, Juan Miguel Pascale, Sandra Lopez-Verges, Alexander A Martinez, on behalf of Gorgas COVID19 team and Panama COVID19 laboratory network

## Abstract

**Background:** With more than 50000 accumulated cases, Panama has one of the highest incidences of SARS-CoV-2 in Central America, despite the fast implementation of disease control strategies. We investigated the early transmission patterns of the virus and the outcomes of mitigation measures in the country.

**Methods:** We collected information from epidemiological surveillance, including contact tracing, and genetic data from SARS-CoV-2 whole genomes, of the first five weeks of the outbreak. These data were used to estimate the exponential growth rate, doubling time and the time-varying effective reproductive number (Rt) using date of symptom onset in a Bayesian framework. The time of most recent ancestor for the introduced and circulating lineages was estimated by Bayesian analysis.

**Findings:** A total of 4210 subjects were SARS-CoV-2 positive during the period evaluated, of them we sequenced 313 cases, detecting the circulation of 10 SARS-CoV-2 lineages. Whole genomes analysis identified the local transmission of one cryptic lineage as early as 2 weeks before it was detected by surveillance systems. Analysis of transmission dynamics showed that lockdown reduced Rt and increased the doubling time, however, these measures did not stop the circulation of this lineage in the country.

**Interpretation:** These results demonstrate the value of epidemiological modeling and genome surveillance to assess mitigation strategies. At the same time, an active search for cryptic transmission clusters is crucial to interrupt local transmission of SARS-CoV-2 in a region.

**Funding:** Ministry of Health, Contribution from private donors and Secretaría Nacional de Ciencia y Tecnología.

**Research in Context:** *Evidence before this study:* In May 2020, we searched for published studies in PubMed and web of Science related to genetic variability and dynamics of SARS-CoV-2 transmission in Latin America, and there was none. On July 2020, there was one study of this type on SARS-CoV-2 transmission in Brazil and none in Central America. We were particularly interested in SARS-CoV-2 cryptic transmission that could allow the virus spread through locals without being detected by respiratory health system surveillance, and no publication was reported. On July 2020, seven papers (five in preprint) were about SARS-CoV-2 cryptic transmission, one in China, another in UK and five in the US. None in Central America. All of them showed the importance of genomic surveillance to detect different lineage introductions, cryptic transmission and its role in early spread in a region or in health-care setting.

*Added value of this study:* We integrate data collected from tested individual during national surveillance of COVID19 suspected cases or contact of cases, as part of the National COVID19 Laboratory network. This data was used to estimate epidemiological parameters of the outbreak as well as the effect of mitigation measures on the epidemic dynamic. We sequence the whole genome of SARS-COV-2 of 7.4% of RT-PCR confirmed cases at the national level, and with phylogenetic analysis we identified SARS-CoV-2 lineages introduced in the country and estimate date of their introductions. Epidemiological and genetic data was compared and we observed the cryptic transmission of one introduced lineage and the rise of a local lineage that was not detected by the active contact tracing implemented by the health system surveillance. This cryptic lineage could explain the fact that early implementation measures decreased the transmission rate and the increased the doubling time, however they were not able to eliminate totally the virus spread.

*Implications of all the available evidence:* This is the first study that analyzed the epidemiology and transmission dynamics of the early COVID19 epidemic in a Central American country using both epidemiological and genomic surveillance. Our findings suggest that strict containment measures and movement restrictions in Panama might have contributed to decrease the early spread of the virus, but that cryptic local transmission allowed a continual basal virus diffusion that could explain, in part, the high incidence of cases in the country. More broadly, our findings are crucial to inform intervention policy in real-time, for countries in similar situations and the importance of constant monitoring of SARS-CoV-2 lineages to understand its transmission in a region.

## Introduction

A severe respiratory disease called Coronavirus Disease 19 (COVID-19) emerged in early December 2019, in Wuhan City, Capital of Hubei province in China^1^. The etiologic agent was designated as SARS-CoV-2, a single strand, positive sense enveloped RNA virus of the genus betacoronavirus, family Coronaviridae^2^. After several zoonotic viral spillover events^2–4^, epidemiological investigations confirmed human-to-human transmission of SARS-CoV-2 among the Wuhan population^5^.

By January 30^th^ 2020, COVID-19 was declared a public health emergency of international concern by the World Health Organization (WHO), and by March 11, COVID-19 was declared a pandemic. Previous analysis of transmission dynamics suggested that SARS-CoV-2 is highly contagious and transmissible among the human population, initial calculations estimated that the basic reproductive number (R_0_) was approximately 2.2^1,2,6^. However, others estimates indicated that the R_0_ could be higher^7^ with a significant role of asymptomatic carriers^8^. As of July 21, SARS-CoV-2 has spread to 188 countries with around 14 763 911 confirmed cases and 611 322 fatalities^9^. To characterize SARS-CoV-2 local transmission across the globe^10^, the few mutations accumulated to date in the viral genome^11–13^ have allowed the characterization of different lineages or clades (A and B)^14^.

In Latin America, SARS-CoV-2 was first reported on February 25^th^, in Brazil^15^. In Panama, the first COVID-19 confirmed case was on March 9^th^ in an inbound traveler returning from Spain. However, on March 7^th^ a fatal case of severe respiratory disease was identified as a COVID-19 suspected case, a tissue sample was collected the 8th and laboratory confirmation was performed the next day. Although Panama rapidly implemented disease control strategies, in Latin America it is among the countries with the highest cumulative incidence rate and fatalities^16^. We hypothesized that the high incidence of COVID-19 observed in Panama was associated with SARS-CoV-2 cryptic transmission and early spread, despite the early laboratory analysis of suspected cases and the rapid implementation of disease control strategies. In this article, we gather data on epidemiological surveillance, contact tracing, and genetics from whole genomes during the first five weeks of the outbreak, to evaluate the dynamics of the introduction and spread of SARS-CoV-2 in Panama.

## Materials and Methods

### Bioethics Permit

The protocol EC-CNBI-202-04-46 was approved by the National Committee on Bioethics of Research of Panama to use de-identified epidemiological data to analyze SARS-CoV-2 transmission and spread, as well as to sequence the complete genome of SARS-CoV-2 from Gorgas Memorial Institute of Health Studies (GMI)’s confirmed cases.

### COVID-19 Surveillance system

The surveillance program for COVID-19 was implemented by the Panama Ministry of Health (MoH) on January 20th, 2020. Suspected cases were actively sought at international airports using the WHO/PAHO case definitions and recommendations^16^. In this first stage, suspected cases were defined as presenting Influenza Like Illness (ILI) and severe acute respiratory infections (SARI) symptoms and signs, as well as coming from China. Later, on March 4^th^, the list of countries was expanded to include Italy, Iran, South Korea, and Japan. Travelers from countries with confirmed circulation of SARS-CoV-2 were isolated at home or hotel-quarantine for 14 days and monitored for onset of symptoms. Patients that met the criteria of suspected cases were then confirmed via laboratory diagnosis. For confirmed cases, contact tracing was performed by the National Epidemiology Department of the MoH, and cases and contacts were placed under quarantine. Clinical evaluation and temperature measurement for travelers and contacts were done every day; if symptoms developed, nasopharyngeal or oropharyngeal swabs were taken by the rapid response teams of MoH and sent to the GMI for laboratory diagnosis. Details of COVID-19 surveillance system are described in the Supplementary Information.

### Laboratory diagnosis

Nasopharyngeal and oropharyngeal swabs samples were submitted to the GMI for laboratory confirmation since January 23rd and to additional laboratories from the National COVID-19 Laboratory network since March 16th. Molecular diagnosis was performed first using a generic coronavirus RT-PCR and then specific Real Time RT-PCR for SARS-CoV-2^17^ as described in the Supplementary Information.

### Epidemiological Investigation

Epidemiological data on suspected cases and their contacts were recorded by physicians when nasopharyngeal and oropharyngeal swabs were taken, using a standardized epidemiological form. Data entry was independently undertaken by the National COVID-19 Laboratory network, and then checked by the National Department of Epidemiology of MoH to confirm the accuracy of the information. A dataset of daily incidence based on date of symptom onset was created. The first 61 days of COVID-19 epidemic in Panama from February 15 to April 16 were analyzed. More details are given in the Supplementary Information.

### Epidemic parameter estimation

Early transmission dynamics of COVID-19 in Panama were evaluated estimating the daily growth rate, doubling time, basic and effective reproductive numbers. We estimated the basic reproductive number (R_0_) using the time series of confirmed cases, an early R_0_ with Likelihood-based estimation using a branching process: This estimation follows Poisson likelihood standards^18^. A serial interval mean of 4·7 days and standard deviation of 2·9 was used for the estimation^6^. The time-varying effective reproductive number (R_t_) was estimated in a Bayesian framework since the cumulative number of cases reached 25 cases as previous decribed^6^, serial interval with 95 % credible intervals (95% Cl), using the EpiEstim packaged implemented in R^18^. Details are provided in Supplementary Information.

### SARS-CoV-2 Sequencing

Reverse transcription and multiplex PCR were performed according to ARTIC Network protocol (https://artic.network/), with modifications. Amplicons generated were pooled and prepared for Illumina Sequencing with the Nextera XT library preparation protocol. All samples were sequenced with MiSeq V2 reagent kits for 500 cycles. Reads obtained were aligned with bowtie-2 and consensus genome was generated with Quasitools-Hydra. Details are provided in Supplementary Information.

### Genome Analysis

To evaluate virus lineage distribution in Panama, lineages were classified using Pangolin^14^, and their distribution was assessed per geographic region of origin of the sample and epidemiological information. Time-trees were estimated using BEAST v1.10.4^19^ and BEAGLE v3^20^ to enhance computational speed. Posterior estimates of most recent common ancestor (tMRCA) of each locally transmitted lineage was obtained as described in Supplementary Information.

### Control measures in Latin America

Reports from the WHO, the Organization for Economic Co-operation and Development (OECD) and official communications from the ministries of health were used to obtain data on control and mitigation strategies at the country level to reconstruct a timeline representing the strategies adopted by Latin American countries. Only when no official information was found, press articles from local newspapers were consulted, as described in Supplementary Information.

## Results

### Early epidemic dynamics in Panama

Epidemiological surveillance began on January 23 and, by April 16, a total of 18 559 suspected cases of COVID-19 had been investigated. Of these, 4 210 were confirmed as SARS-CoV-2 infections at that date. On March 7th, the National Surveillance system detected a fatal case in which the SARS-CoV-2 infection was confirmed on March 9th. This case was not associated with previous travel history outside the country. The first case detected by active surveillance at international airports, came from Spain, had mild disease and was confirmed on March 9th. Most imported cases were associated with travel from Europe, North America, and one from China.

Epidemiological investigation confirmed that the date of onset of symptoms of one local case in our study goes back as early as February 15 (Figure 1A). Most local cases had mild disease (Figure 1B). By April 16, a total of 341 hospitalized cases (77 were hospitalized at diagnosis confirmation) and 116 fatalities (31 were dead at the diagnosis confirmation) were reported in Panama (Figure 1C and 1D). The highest proportion of confirmed cases were observed in the age group 20-59 years (Supplementary Figure 1A). A higher proportion of females were tested while more males were positive (Supplementary Figure 1B). A rapid growth rate of 0·13 cases/per day (Figure 2A) and a short doubling time was also observed during the early stages of the epidemic, with an increasing doubling time over the analyzed interval (Figure 2B). We estimated an early epidemic potential of transmission of SARS-CoV-2 in Panama of R_0_= 2·22 (95% CI, 2·08 - 2·37).

**Figure 1.**
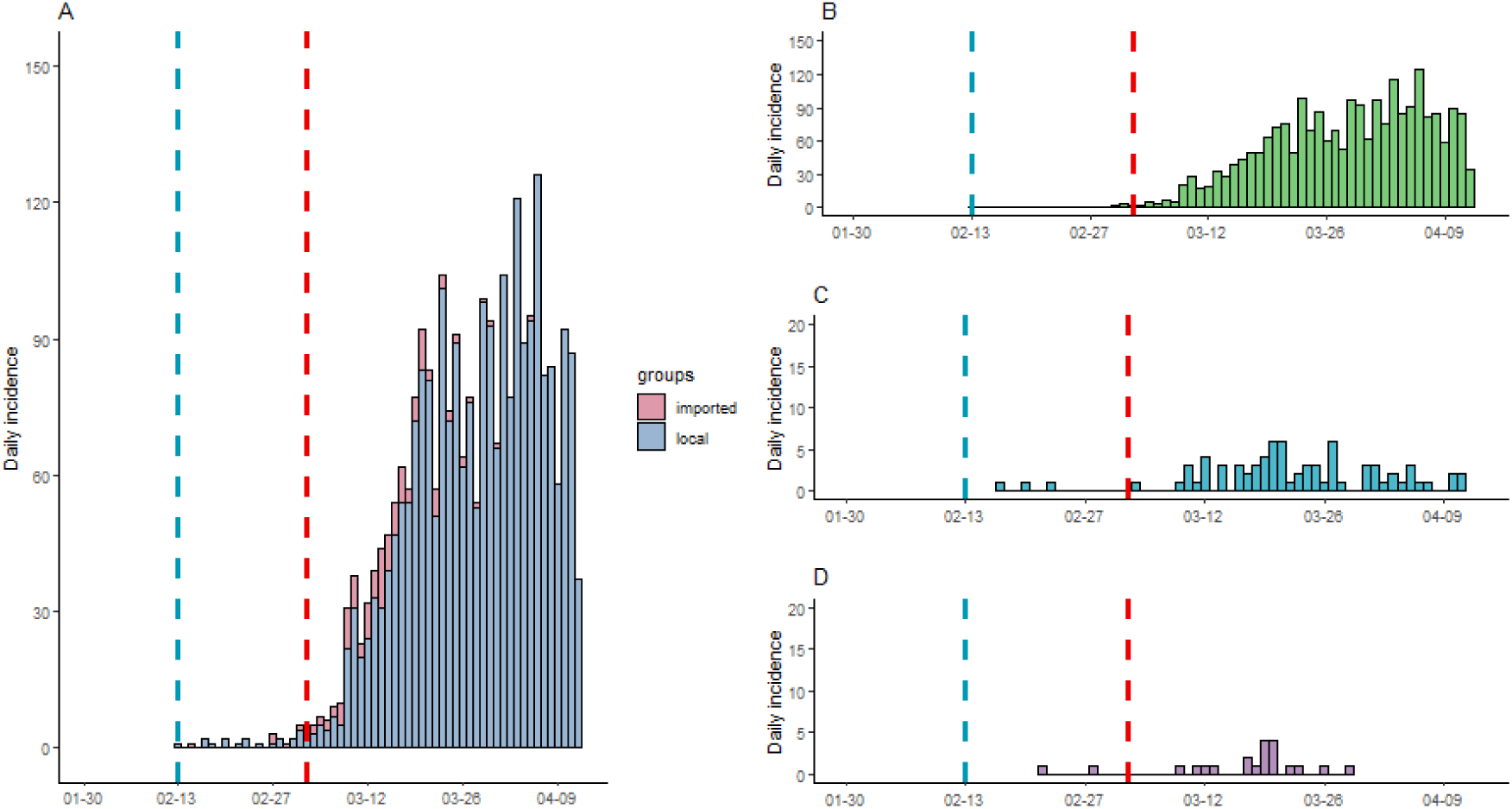
Epidemic Curve of confirmed SARS-CoV-2 infections. Daily incidence of: **A)** imported and local confirmed infections, **B)** ambulatory cases (asymptomatic, pre-symptomatic, mild outpatient cases), **C)** hospitalized cases and **D)** fatal cases, with symptom onset from February 13^th^ to April 13^th^. In all four charts, the y axis represents the daily incidence and the x axis represents the date of symptom onset for each reported case. The blue vertical dashed lines represent the first recorded onset of symptoms and vertical red dashed lines the date of first confirmed case by the surveillance system.

**Figure 2.**
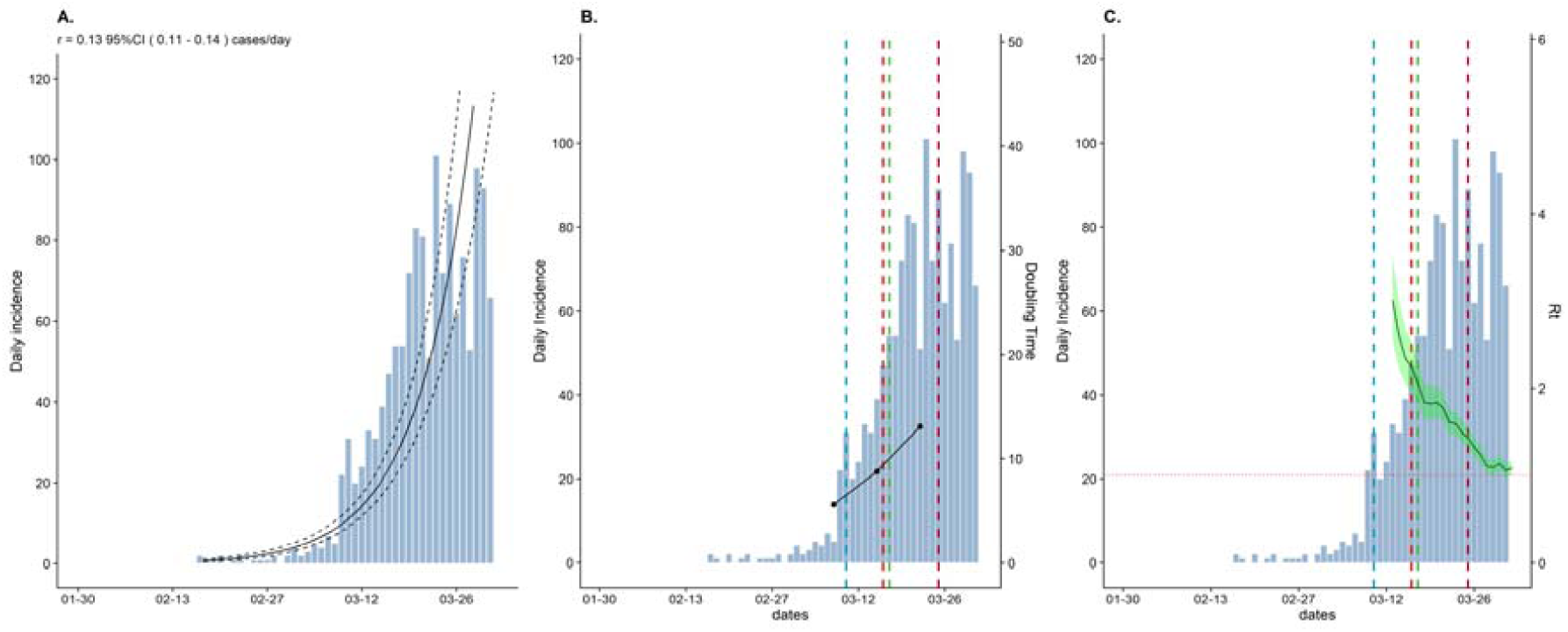
Epidemiologic parameters: Number of COVID-19 daily confirmed infections (left axis and bars) overlaid with estimates of **A)** Fitted exponential growth in cases daily growth rate, **B)** Doubling time and **C**) Time-varying effective reproduction number *R*, for a time frame of 45 days (x axis). For C), green shaded areas show 95% credible intervals around the median estimated *R*. The threshold value *R*_*t*_ = 1 is indicated by the horizontal red dashed line. For both B) and C), the vertical blue dashed line represents the schools’ closure, the vertical lines red and green represent night curfews and restrictions of movement respectively, and the purple dashed line the 24-hour curfew.

Panama detected the first SARS-CoV-2 infection one month after Brazil^15^, being the 11^th^ country with confirmed cases in Latin America (Supplementary Figure 2). School closures were implemented the day after first case confirmation, social distance measures six days later, and stay-at-home 24-hour curfew 14 days later. Compared to other countries in the Latin American region, Panama was among those that rapidly implemented control strategies after its first case (Supplementary Figure 2). The effective reproduction number since the accumulated 25 cases was R_t_= 3·01 (95% Crl: 2·54 - 3·52) and rapidly dropped over the course of 17 days to Rt= 1·08 (95% Crl: 1·00 −1·17) (Figure 2C, Supplementary Table 1). Until April 16, Panama was the country from Central America with the highest cumulative incidence of cases and fatality (Supplementary Figure 3).

### SARS-CoV-2 lineage distribution in Panama

To characterize the distribution of genetic lineages in Panama, we generated 313 SARS-CoV-2 sequences, which represents 7.4 % of the total confirmed cases by April 15^th^ (Supplementary Figure 4A). Analyzed sequences were obtained from patients with symptom onset between February 15 and April 14. The genome coverage of the analyzed sequences increased as the real-time PCR cycle threshold (CT) value of the sample decreased, with the best coverage obtained at CT values lower than 25 (Supplementary Figure 4B). Lineage analysis showed the circulation of ten previous reported lineages (Figure 3A, Supplementary Figure 4). The most successfully distributed lineage was A.2 (71.2 %), followed by B.1 (16.7%), and A.1 (3.5%). Lineages A.3, B, B.1.5, corresponded to early detected cases, most of them from returning travelers (squared tips in Figure 3A and black outlined circles in Supplementary Figure 4). Lineage A.2 was found in subjects with an epidemiologic link to cases related with a school outbreak (1.8%) where the first local transmission was detected, and in police officers (6.3%). Additionally, a group of 170 sequences which had an C to T nucleotide substitution at position 12815 emerged as a sister clade from lineage of A.2 (Figure 4A). These sequences, indicated a substantial local transmission as they corresponded to 54·3% of the sequenced cases in the country. Estimates of the tMRCA of the Panamanian sequences place the median age of A.1, A.3, B.1 between two weeks range before the first reported COVID-19 case, which agrees well with the time of onset of symptoms observed in the data (Figure 1A, Figure 3A), however the A.2, had the oldest median tMRCA corresponding to early February (Figure 3A).

**Figure 3.**
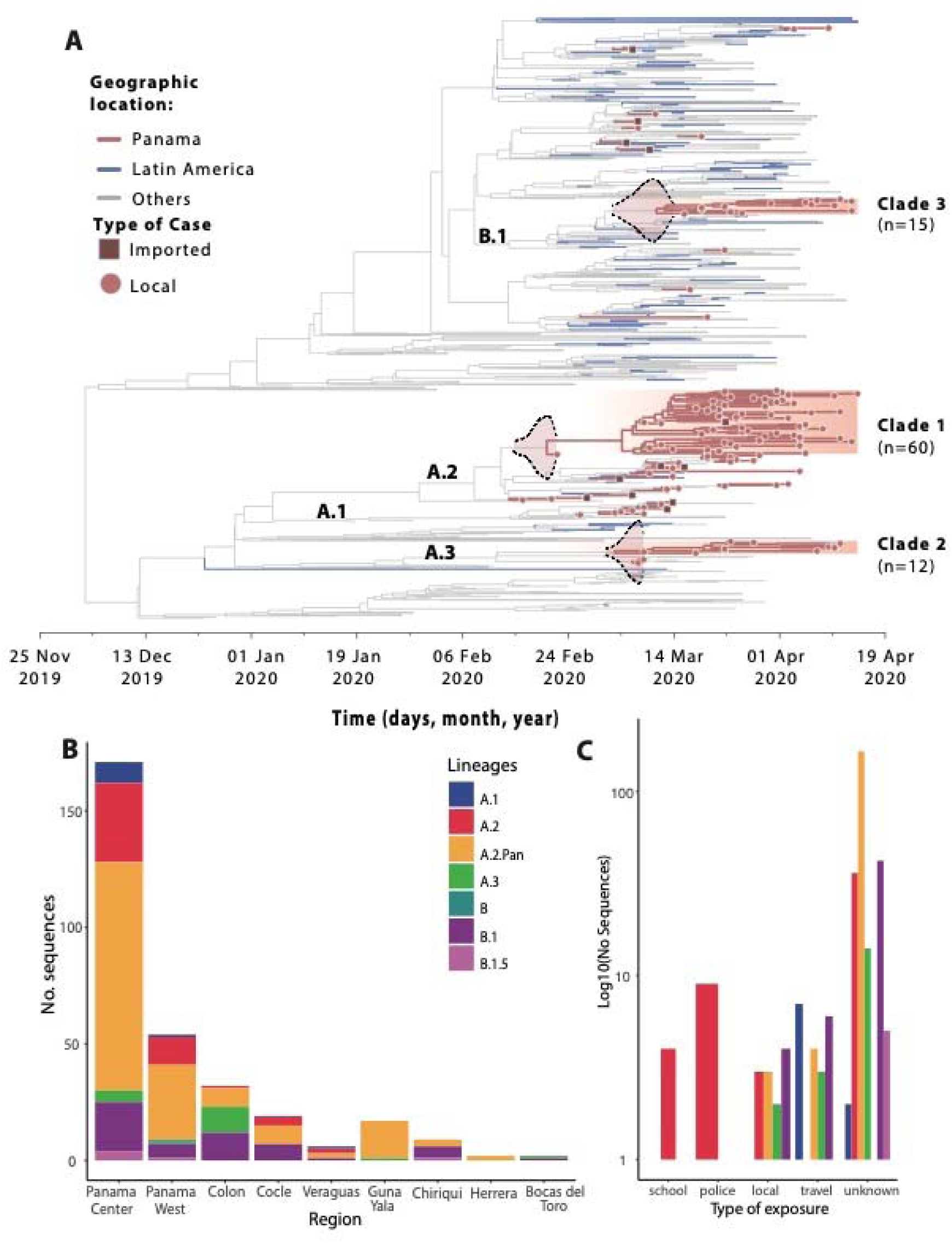
Time-tree resolution, geographic and epidemiological characteristics of the SARS-CoV-2 genomes obtained. **A)** Bayesian maximum clade credibility tree of 1261 SARS-CoV-2 sequences, 133 from Panama (red), 91 from Latin America (blue), 904 from other location (grey), tips with box shapes represent samples that have a clear epidemiological travel link. Posterior density estimates of time of the most recent common ancestor of each lineage with local transmission are show in their branches. **B)** Distribution of the lineages among regions in the country. **C)** Distribution of the lineages by type of exposure detected by the surveillance system.

**Figure 4.**
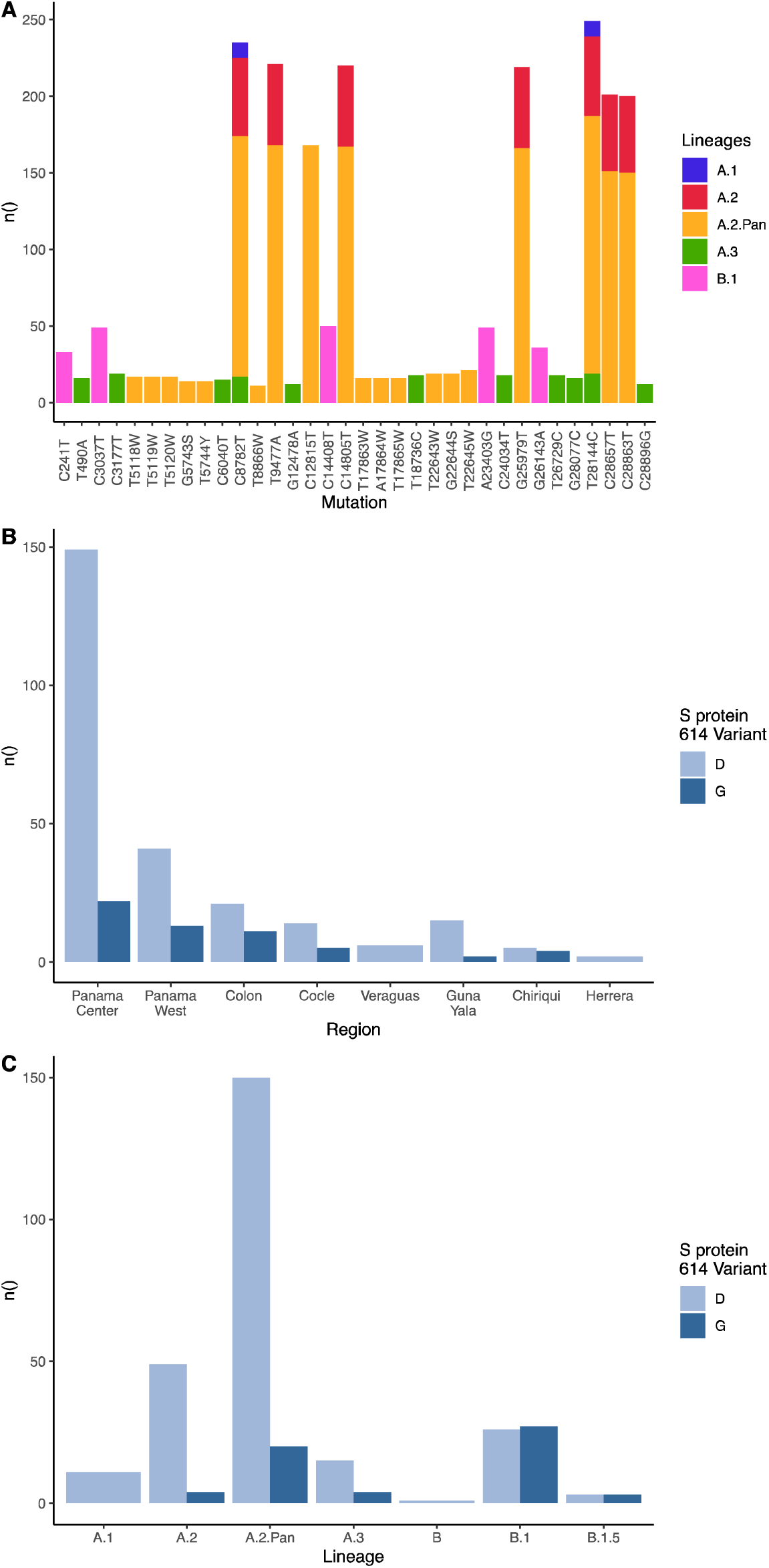
Mutation profile of the sequences obtained in the study. **A)** Frequency of single nucleotide polymorphism in the genome (position according to MN908947) for all analyzed sequences (n=313) in the study. Distribution of S protein variants (D614 or G614) **B)** in different regions of the country, and **C)** among lineages.

There is no structured spatial distribution of the lineages across the country, however the locally transmitted A.2 linage was found in all regions. Additionally, Center Panama and West Panama (regions that correspond to Panama City and its suburbs), had a more diverse lineage distribution (Figure 3B). Using active contact tracing of positive cases at the beginning of the outbreak, the National Epidemiology Department of MoH defined 30 epidemiological clusters (supplementary table S2), linked to a school, police officers and local outbreaks. Most sequenced cases were not classified inside these epidemiological clusters (unknown category in Figure 3C) and had a predominance of lineage A.2.

The previously described spike glycoprotein variants D614 and G614^21,22^ were co-circulating early in the epidemic among all the regions analyzed, but G614 variant was predominant in 18.8% of the sequences analyzed (Figure 4B). Both variants were also distributed among lineages, however D614 was highly predominant in A.1 and A.2. (Figure 4C).

## Discussion

Panama has the most confirmed SARS-CoV-2 infections and associated fatalities in Central America, although control strategies were rapidly implemented at the beginning of the outbreak. The reasons for this pattern are unclear, but they may include, 1) cryptic circulation, 2) rapid early transmission dynamics and 3) actively search of cases and their contacts, combined with strong molecular capacity of testing in the country.

Epidemiological investigation showed early local transmission events as February 23^th^, based on reports of symptoms onset of the first detected local case on March 9^th^. The potential source of this infection dated back an onset of symptoms on February 15^th^. Taken together the epidemiological evidence, along with genetic analysis of SARS-CoV-2, confirm cryptic circulation in Panama with an interval window of introduction between late January and early February.

A high early median transmission potential of SARS-CoV-2-was estimated in R_0_= 2.22 (2.08-2.37), like those early estimated reported in China and Europe^23,24^. In addition, the initial time-variant reproduction number R_t_=3.01 (2.54-3.51), estimated since the incidence reached 25 cases, also suggest a high early transmission dynamic. However, the R_t_ rapidly dropped after implementation of control strategies.

The genetic analysis of the diverse introductions of lineages and the dynamics of their transmission support these epidemiological observations. At the beginning of the outbreak, we detected the introduction of ten lineages, and from them, five were still co-circulating in the country in mid-April. The numbers of detected lineages could be underestimated because we did not sequence all positive cases, thus there is a possibility of not-detecting uncommon lineages. Most of the lineages associated with imported cases (A.1, A.3, B, B.1, B.2.1) were detected by surveillance of suspected cases and their contacts through active contact tracing. However, this did not happen with the cases associated to the lineage A.2. According to our phylogenetic dating analysis, this lineage circulated in the country at least three weeks before it was first detected. This lineage was detected among the school and the police officers clusters, and most of the cases belonging to these groups had their onset of symptoms between February 15^th^ and March 15^th^, including the first detected fatal case. The failure to detect and control the spread of these clusters gave enough time to A.2 lineage to acquire a synonymous mutation that was fixed in the sequences analyzed and became the A.2 lineage locally transmitted reported in this study. This descendent lineage (A.2.Pan) was detected in all regions of the country and was not associated with any epidemic transmission cluster isolated by the public health systems. Moreover, our observations suggest that in Panama we had a local transmission of lineage A.2.Pan one week before respiratory disease surveillance detected it. Once this lineage was detected, active contact tracing and testing activities promptly confirmed more cases. More extensive sampling should be implemented in the country following well-established pipelines as suggested previously.^25^

When analyzing the different lineages, the most common mutations (C8782T and T28144C) observed in the analyzed sequences have been previously reported in South America, United States, Europe and China^26,27^, whereas the observed mutations T9477A and C28657T were reported in South America^28^. These mutations were not associated with a specific lineage nor with specific transmission or pathogenic effects. The C12815T mutation detected only in the A.2.Pan lineage has not been reported to date.

The D614G variant was detected in all lineages circulating in Panama during the beginning of SARS-CoV-2 outbreak. This variant was also reported in Europe, Mexico, Brazil, and Wuhan since early February 2020. It has been related to more efficient virus transmission,^29^ but with our data do not support the predominance of this variant in the circulating lineages.

The pattern of cryptic transmission in Panama was also observed in other countries which also experienced early and rapid dissemination of SARS-CoV-2.^27,30^ We suspect that the tight classification of cases at the beginning of the surveillance in January, which focused mainly in symptomatic China travelers, precluded the opportunity to detect these early importations from Europe and USA. Indeed, in these regions the virus was already circulating at that time.^23,27,30^ Our forecast of growth rate and R_0_ showed that mitigation measures undertaken by MoH helped to reduce the number of new cases of SARS-CoV-2. Implementation of epidemiological control measurements such as active contact tracing and isolation, social distancing and region-based quarantine where active transmission clusters are found, would help to effectively control the spread of SARS-CoV-2 as suggested by results obtained in other countries.^24^ To implement and maintain these measures until the outbreak is under control, it is important to improve the communication strategy to provide the public with the appropriate advice to better prevent infection and subsequent transmission. Improvements in the surveillance and tracing, data collection and digitalization systems should increase the capacity of the country to respond to any new transmission waves.

## Data Availability

Data and materials availability: All data, and materials used in the analysis are available on our GitHub repository available upon acceptance.

## Acknowledgments

Thanks to Gorgas’ administration, general services for their support and extra-work during the COVID19 response. Thanks to all the Health institutions from MINSA, CSS and private hospitals. AAM, JMP, SLV, LEA, JMP are members of the Sistema Nacional de Investigación (SNI) del SENACYT, Panamá.

